# Differential DNA Methylation and Delirium After Anesthesia and Surgery

**DOI:** 10.64898/2026.06.12.26355544

**Authors:** Kirk J. Hogan, Miles Berger, Sydney P. Kolstad, Andy Madrid, Bethany Hsia, Mary C. Wright, Michael Devinney, Melody R. Smith, Reid S. Alisch

## Abstract

**Background:** DNA methylation is an epigenetic modification that regulates gene expression in response to environmental exposures. We measured differential DNA methylation levels in blood before after general anesthesia and surgery in participants with and without postoperative delirium (POD) and postoperative neurocognitive disorder (PNCD).

**Methods:** Blood sampling, delirium assessment and cognitive testing were prospectively performed at baseline before non-cardiac, non-neurologic surgery, and at 24 hours (24h) and 6 weeks (6wk) thereafter in 94 participants comprising 13 with POD and 81 without POD, and 40 with PNCD and 54 without PNCD 6wk after surgery who were matched for age and sex in the INTUIT and MADCO cohorts. DNA methylation was assessed using the Illumina Infinium MethylationEPIC Beadchip.

**Results:** 132 differentially methylated positions (DMPs) annotated to 198 differentially methylated genes (DMGs) were identified in 94 participants 24h after surgery compared to baseline with a local false discovery rate (LFDR) <0.05 including *CHRNB1*, *LGALS1*, *SMAD4*, *RYR2*, *CHST11*, *CDC25B*, *OBSCN*, *ABHD16A*. No DMPs were identified between samples collected at baseline compared to 6wk after surgery. In baseline samples, 8 DMPs annotated to 12 DMGs were identified between participants who did and did not develop POD including *MBTD1, BID, PPAN, ANGPTL6, PHF21B,* and *RBM5*. In 24h samples, 87 DMPs annotated to 91 DMGs were identified between participants with and without POD including *CLEC19A, FILIP1, ERICH1, PSENEN, SLC6A3,* and *TMEM196.* In 6wk samples, 1 DMP annotated to *FILIP1* and *LOC124901509* in participants with and without POD. No DMPs in baseline, 24h or 6wk blood samples were identified between patients with and without PNCD at 6k after surgery.

**Conclusions:** Differential DNA methylation levels are present throughout the genome 24h after anesthesia and surgery. Differential DNA methylation levels before surgery and at 24h after surgery distinguishes patients with and without POD. Differential DNA methylation levels were not identified between baseline and 6wk after surgery in the entire cohort, or between patients with and without PNCD at 6 weeks.

## Introduction

Over 10% of patients experience postoperative delirium (POD) characterized by fluctuating confusion, disorientation, impaired perception and attention deficits that arise within days after surgery. (Inouye, 2014, Maracantonio, 2012, Schenning, 2015) POD is associated with higher rates of postoperative complications, longer hospital stays, and increased progression to dementia and mortality. (Marcantonio, 1994, Witlox, 2010, Mahanna-Gabrielli, 2019, Inouye, 2016, Saczynski, 2012, Raats, 2016) Postoperative neurocognitive disorder (PNCD) (Evered, 2018) is characterized by an objectively measured 1-2 standard deviation (S.D.) decrement in cognitive performance from before surgery to 1-12 months after surgery assessed by neuropsychological testing combined with a subjective complaint of impaired cognition. (Needham, 2017, Daiello, 2019, Moller, 1998, Paredes, 2016, McDonagh, 2010, Deo, 2011, Oughli, 2018, Mahanna-Gabrielli, 2018, Oughli, 2018, Peden, 2021) Cognitive decline after surgery (also referred to as postoperative cognitive dysfunction (POCD) in older literature) is associated with persistent cognitive impairment up to 7 years after surgery, increased loss of independence and reduced quality of life, higher health care costs and increased morbidity and mortality. (Abildstrom, 2000, Berger, 2015, Monk, 2008, Steinmetz, 2009) The etiologies of POD and PNCD are poorly understood. No biomarkers of POD and PNCD risk, onset and progression have been introduced into practice.

CpG sites within the human genome comprise a locus in which a cytosine nucleotide (C) is followed by a guanine nucleotide (G) on the same DNA strand linked by a phosphate group. Covalent addition of a methyl group at position 5 of the C nucleotide generates 5-methylcytosine (5mC). The methylation status of CpG dinucleotides is important because 5mC participates in regulation and coordination of gene expression in the human genome. (Mattei, 2022) Fluctuations in 5mC levels across the genome occur over the lifespan in association with environmental exposures and changes in cognitive status arising from neurodegenerative disorders. (Sanchez-Mut, 2016, Younesian. 2022, Day, 2013) CpGs with differential methylation in the presence and absence of a trait or exposure are identified as differentially methylated positions (DMPs). A single DMP may appear in more than one differentially methylated gene (DMG) in keeping with local gene structure and overlap. Using the Illumina Infinium MethylationEPIC Beadchip we’ve reported multiple DMPs in blood DNA that distinguish patients with and without Alzheimer’s disease in numerous DMGs and in loci between genes, thereby underscoring the potential use of blood DMPs to serve as biomarkers of changes in gene expression and cognition after anesthesia and surgery. (Madrid, 2018)

Methods to detect differential DNA methylation levels have not previously been applied to blood samples collected from patients at the time of neuropsychological testing for POD and PNCD, or at a time point after immediate recovery from anesthesia and surgery. We used the Illumina Infinium MethylationEPIC BeadChip to compare DNA methylation levels in blood before non-cardiac, non-neurologic surgery and general anesthesia at baseline, and at 24 hours (24h) and 6 weeks (6wk) thereafter from cognitively unimpaired participants in two prospective perioperative cognition cohorts: “Investigating Neuroinflammation Underlying Postoperative Brain Connectivity Changes (INTUIT)” (Berger, 2019) and “Markers of Alzheimer’s Disease and Neurocognitive Outcomes after Perioperative Care (MADCO-PC)” (Berger, 2022)

## Methods

### Participants

Samples and data were obtained from the Investigating Neuroinflammation Underlying Postoperative Cognitive Dysfunction study (INTUIT; NCT03273335), (Berger, 2019, Acker, 2024), and from the Markers of Alzheimer’s Disease and Neurocognitive Outcomes after Perioperative Care study (MADCO-PC; NCT01993836). (Browndyke, 2021, Berger, 2022) The INTUIT and MADCO-PC prospective observational studies were approved by the Duke Institutional Review Board. All participants provided written informed consent before enrollment. INTUIT enrolled 201 surgical participants 60 years of age and older undergoing non-cardiac, non-neurologic surgery. MADCO-PC enrolled 140 surgical participants over 60 years of age and older undergoing non-cardiac, non-neurologic surgery. Participants were scheduled for elective procedures of at least 2 hours duration under general anesthesia with a planned overnight hospital stay of at least one night at Duke University Hospital or Duke Regional Hospital. No study restrictions were placed on coincident use of regional or neuraxial blocks (*e.g.,* spinal or epidural anesthesia), adjunct anesthetic drugs or surgical technique. Participant characteristics and clinical variables were recorded from the electronic medical record. Participants were administered the Duke Subjective Cognitive Function and Duke Activity Status questionnaires preoperatively. (Hlatky, 1989) Charlson and Elixhauser Van Walraven comorbidity scores and apolipoprotein E4 status were recorded. (Charlson, 1994, Elixhauser, 1998, Quan, 2005, Brownkyke, 2021) Surgery type was classified based on the operative surgical service. Length of stay was ascertained from the electronic medical record. Samples were selected such that participants in the POD and No-POD and in the PNCD and No-PNCD cohorts were matched for age, sex, ancestry, level of education and baseline cognitive status.

### Delirium Assessment

Delirium was assessed twice daily with the 3-minute diagnostic Confusion Assessment Method (CAM) in INTUIT, (Marcantonio, 2014), and with the 3D-CAM with a skip pattern in which assessment was stopped if inattention was not present in MADCO-PC. The 3D-CAM and CAM are sensitive and specific for delirium in hospitalized patients when used by trained staff. (Marcantonio, 2014, Inouye, 1990) When a participant was intubated or was otherwise nonverbal at the time of assessment, the Confusion Assessment Method for the Intensive Care Unit (CAM-ICU) method of delirium assessment in intubated/nonverbal patients was used. (Ely, 2001) A validated chart review method was performed to identify delirium missed by research assessments performed at discrete time points. (Inouye, 2005) Participants were defined as having POD as a dichotomous outcome if they had at least one positive bedside CAM assessment (3D-CAM, CAM, or CAM-ICU) or a positive delirium chart review within 7 days after surgery.

### PNCD testing

Preoperative cognitive function was measured with a 14-item test battery including the Wechsler Test of Adult Reading, the Revised Wechsler memory scale and Modified Visual Reproduction test, the Hopkins verbal learning test, the Randt Short Story memory test, the Digit Span test, the Trail Making Test A, the Trail Making Test B, the Digit Symbol test, and the Lafayette Grooved Pegboard Test. (Berger, 2019, Newman, 2001, Browndyke, 2021) The tests generated a total of 14 different scores for factor analysis. The Trail Making Test B was truncated at 300 seconds. The Trail Making Tests were negatively log-transformed such that higher scores indicated better performance in keeping with other test variables. Factor analysis was performed with oblique rotation to generate a 5-factor solution that explained 82% of the variability in test scores. The 5 factors correspond to 5 cognitive domains comprising attention/concentration, structured verbal memory, unstructured verbal memory, visuospatial memory, and executive function. The mean of the 5 cognitive domain scores generated the continuous cognitive index (CCI) score as a measure of baseline cognitive function. (Berger, 2019, Browndyke, 2021) A CCI change from baseline to after surgery quantifies the magnitude of cognitive decline. (Newman, 2001) PNCD as a dichotomous outcome was defined as a one or more standard deviation (S.D.) decrement from before surgery to 6wk after surgery in one or more cognitive domains and at least one subjective complaint.

### DNA extraction and DNA methylation data generation

Whole blood was collected into a 10 ml EDTA tube with plasma removed after centrifugation. The buffy coat was stored at –80°C. Samples were thawed and genomic DNA was extracted using the Gentra Puregene Blood kit following the manufacturer’s protocol (Qiagen, Hilden, Germany). Extracted genomic DNA was resolved on a 1% agarose gel to verify that the DNA was of high molecular weight, and was quantified using QubitTM (Qiagen, Hilden, Germany). Five hundred nanograms of genomic DNA per patient were sent to the University of Illinois at Urbana-Champaign Roy J. Carver Biotechnology Center for sodium bisulfite conversion of unmethylated cytosines to uracil residues using the EZ DNA Methylation-GoldTM kit (Zymo Research, Irvine, CA, USA). The converted DNA was purified and prepared for analysis on the Illumina HuvmanMethylationEPIC BeadChips^TM^ according to manufacturer protocols (Illumina, San Diego, CA, USA). Image analysis, and signal determinations were performed using the GenomeStudio software Methylation Module (Illumina, San Diego, CA, USA). Raw idat intensity files were forwarded to Dr. Alisch’s laboratory via a secure website.

### Illumina HumanMethylationEPIC data preprocessing

Raw idat intensity files were imported to the R environment v4.5.2. Ninety-four participant samples were analyzed at baseline, 24h and 6wk. R package *sesame* was used to read raw intensity files, calculate detection *P*-values for each probe, generate beta-values for each probe, and normalize the dataset using the P-vals Out-Of-Band Array Hybridization (POOBAH) method (Zhou, 2018). Probes were removed from analysis if one sample at most had a detection *P*-value > 0.01, if the probes reported methylation at known SNPs and CpH sites, if the probes were known to be cross-reactive (https://zwdzwd.github.io/InfiniumAnnotation#reference), or if the probes were located on sex chromosomes to provide 621,455 probes from EPIC v.1 (baseline and 6wk) and 749,254 probes from EPIC v.2 (24h) microarrays. After integration of the datasets a total of 558,817 probes were analyzed for differential DNA methylation for within-array version comparisons. To confirm between-array version reproducibility, 9 samples were tested using both EPIC array versions. A differential analysis was computed with array version entered as a fixed effect and individual sample ID entered as a random effect. Probes were determined to be replicable if they were not significantly different (local false discovery rates [LFDR] > 0.05) and their methylation difference was less than 2.5% between array versions. The 558,817 probes provided for within-array comparisons were filtered to provide 437,286 replicable probes for between-array version comparisons. Cell type proportions were calculated from the filtered probes using R package *meffil*. (Min, 2018)

### Statistical analysis

The R package *lmerTest* (Kuznetsoava, 2017) was used to apply linear mixed effects models to filtered probes to test differential methylation between baseline, 24h and 6wk time points. Sample date, sex, age, ancestry, and estimated cell proportions were entered as fixed effects, with individual sample ID entered as a random effect. R package *emmeans* was used to contrast timepoints. (Lenth, 2025) The empirical distribution of the 558,817 and 437,286 *P*-values deviated from uniform distribution with an enrichment around small *P*-values when comparing baseline or 6wk time points to 24h time points with inflation of *P-*values. Accordingly, test statistics were used to model the empirical null and to calculate local false discovery rates (LFDR) with R package *fdrtool* to mitigate over-inflation of *P*-values and to correct for multiple testing. (Stimmer, 2008) CpG loci with a LFDR < 0.05 and a mean differential methylation between time intervals > 2.5 % were identified as differentially methylated positions (DMPs). CpG loci were annotated to genes with R package annotatr. DNA sequences spanning 5 kilobases (kb) 5′ of a transcription start site (TSS) to 200 base pairs (bp) 3′ of a transcription termination site (TTS) were defined as genes (Cavalcante, 2017). A differentially methylated gene (DMG) comprises at least one DMP, and a DMP may occupy more than one DMG.

## Results

### Participants

94 surgical patients matched for age (68.62 years (±6.13) and sex (48% female) were ascertained in the INTUIT and MADCO-PC cohorts for inclusion. (Table 1.) Urologic (26.6%), orthopedic (24.5%) and general surgery (25.5%) procedures were most common. Participants in the POD and No-POD cohort (Table 1) and in the PNCD and No-PNCD cohort (Table 2) were matched for age, sex, ancestry, level of education and baseline cognitive status.

**Table 1.** Cohort summary by POD status. Numeric variables summarized by mean (SD) or median [Q1, Q3] categorical variables summarized by count (%).

|  | Overall<br>(N=94) | No-POD<br>(N=81) | POD<br>(N=13) |
| --- | --- | --- | --- |
| Age | 68.62 (6.13) | 68.06 (6.13) | 72.08 (5.09) |
| Sex (Male) | 49 (52.1%) | 42 (51.9%) | 7 (53.8%) |
| Race |  |  |  |
| Black or African American | 14 (14.9%) | 10 (12.3%) | 4 (30.8%) |
| Caucasian/White | 80 (85.1%) | 71 (87.7%) | 9 (69.2%) |
| BMI | 29.84 (5.40) | 30.01 (5.43) | 28.80 (5.32) |
| SURGICAL_SERVICE |  |  |  |
| Thoracic | 13 (13.8%) | 10 (12.3%) | 3 (23.1%) |
| General Surgery | 24 (25.5%) | 21 (25.9%) | 3 (23.1%) |
| Gynecology | 5 (5.3%) | 5 (6.2%) | 0 (0.0%) |
| Orthopedics | 23 (24.5%) | 20 (24.7%) | 3 (23.1%) |
| Otolaryngology Head and Neck | 3 (3.2%) | 2 (2.5%) | 1 (7.7%) |
| Plastic Surgery | 1 (1.1%) | 1 (1.2%) | 0 (0.0%) |
| Urology | 25 (26.6%) | 22 (27.2%) | 3 (23.1%) |
| ASA |  |  |  |
| 1 | 1 (1.1%) | 1 (1.2%) | 0 (0.0%) |
| 2 | 28 (29.8%) | 26 (32.1%) | 2 (15.4%) |
| 3 | 64 (68.1%) | 54 (66.7%) | 10 (76.9%) |
| 4 | 1 (1.1%) | 0 (0.0%) | 1 (7.7%) |
| Surgery Duration (min) | 132 [91, 185] | 128 [91, 183] | 145 [114, 202] |
| Years of Education | 16 [13, 18] | 16 [14, 18] | 14 [12, 18] |
| BL MMSE | 29 [27, 29] | 29 [28, 29] | 27 [24, 29] |
| History of Lymphoma | 2 (2.1%) | 2 (2.5%) | 0 (0.0%) |
| History of Metastatic Cancer | 1 (1.1%) | 1 (1.2%) | 0 (0.0%) |
| History of Tumor without Metastasis | 32 (34.0%) | 29 (35.8%) | 3 (23.1%) |
| Charlson Comorbidity Index | 4 [2, 5] | 4 [2, 5] | 4 [3, 5] |
| Elixhauser Van Walraven Index | 0 [0, 4] | 0 [0, 4] | 3 [0, 7] |
| DASI Score* | 20 [11, 37] | 21 [11, 38] | 16 [8, 32] |
| APOE4 Carrier | 34 (36.2%) | 28 (34.6%) | 6 (46.2%) |
| Hospital LOS (days) | 1 [1, 3] | 1 [1, 2] | 4 [2, 5] |

\*DASI missing for 2 subjects in the POD group

**Table 2.** Cohort summary overall and 6-week PNCD status. Numeric variables summarized by mean (SD) or median [Q1, Q3] categorical variables summarized by count (%).

|  | Overall<br>(N=94) | No- PNCD<br>(N=54) | Yes PNCD<br>(N=40) |
| --- | --- | --- | --- |
| Age (years) | 68.62 (6.13) | 68.44 (6.27) | 68.85 (6.01) |
| Sex (Male) | 49 (52.1%) | 29 (53.7%) | 20 (50.0%) |
| Race |  |  |  |
| Black or African American | 14 (14.9%) | 7 (13.0%) | 7 (17.5%) |
| Caucasian/White | 80 (85.1%) | 47 (87.0%) | 33 (82.5%) |
| BMI | 29.84 (5.40) | 30.70 (5.26) | 30.68 (5.32) |
| Surgical Service |  |  |  |
| Thoracic | 13 (13.8%) | 10 (18.5%) | 3 (7.5%) |
| General Surgery | 24 (25.5%) | 13 (24.1%) | 11 (27.5%) |
| Gynecology | 5 (5.3%) | 4 (7.4%) | 1 (2.5%) |
| Orthopedics | 23 (24.5%) | 10 (18.5%) | 13 (32.5%) |
| Otolaryngology Head and Neck | 3 (3.2%) | 2 (3.7%) | 1 (2.5%) |
| Plastic Surgery | 1 (1.1%) | 1 (1.9%) | 0 (0.0%) |
| Urology | 25 (26.6%) | 14 (25.9%) | 11 (27.5%) |
| ASA |  |  |  |
| 1 | 1 (1.1%) | 1 (1.9%) | 0 (0.0%) |
| 2 | 28 (29.8%) | 17 (31.5%) | 11 (27.5%) |
| 3 | 64 (68.1%) | 35 (64.8%) | 29 (72.5%) |
| 4 | 1 (1.1%) | 1 (1.9%) | 0 (0.0%) |
| Surgery Duration (min) | 132 [91, 185] | 142 [98, 229] | 122 [90, 156] |
| Years of Education | 16 [13, 18] | 16 [14, 18] | 16 [13, 18] |
| BL MMSE | 29 [27, 29] | 29 [28, 29] | 29 [27, 29] |
| History of Lymphoma | 2 (2.1%) | 1 (1.9%) | 1 (2.5%) |
| History of Metastatic Cancer | 1 (1.1%) | 1 (1.9%) | 0 (0.0%) |
| History of Tumor without Metastasis | 32 (34.0%) | 19 (35.2%) | 13 (32.5%) |
| Charlson Comorbidity Index | 4 [2, 5] | 4 [2, 5] | 4 [3, 5] |
| Elixhauser Van Walraven Index | 0 [0, 4] | 1 [0, 5] | 0 [-1, 3] |
| DASI Score* | 20 [11, 37] | 21 [10, 38] | 19 [11, 37] |
| APOE4 Carrier | 34 (36.2%) | 21 (38.9%) | 13 (32.5%) |
| Hospital LOS (days) | 1 [1, 3] | 2 [1, 4] | 1 [1, 2] |

\*BL DASI missing for 2 subjects in the No PNCD group

### Differential DNA methylation 24 hours and 6 weeks after anesthesia and surgery

132 differentially methylated positions (DMPs) in 198 differentially methylated genes (DMGs) were identified in 94 participants 24h after surgery compared to baseline (local false discovery rate (LFDR) <0.05 including *CHRNB1*, *LGALS1*, *SMAD4*, *RYR2*, *CHST11*, *CDC25B*, *OBSCN*, *ABHD16A* (Fig. 1a, Supplemental Table 1). DMPs at 6wk compared to 24h (Fig. 1b, Supplemental Table 2) 24h returned to baseline methylation levels at 6wk (Fig. 1c). No DMPs were identified between samples collected at baseline compared to samples collected 6wk after surgery. Every chromosome had at least one DMP between samples collected at baseline and at 24h after anesthesia and surgery. These data indicate that anesthesia and surgery cause extensive and widespread differential DNA methylation between baseline and 24h that reverts to baseline levels 6wk thereafter.

**Fig. 1.**
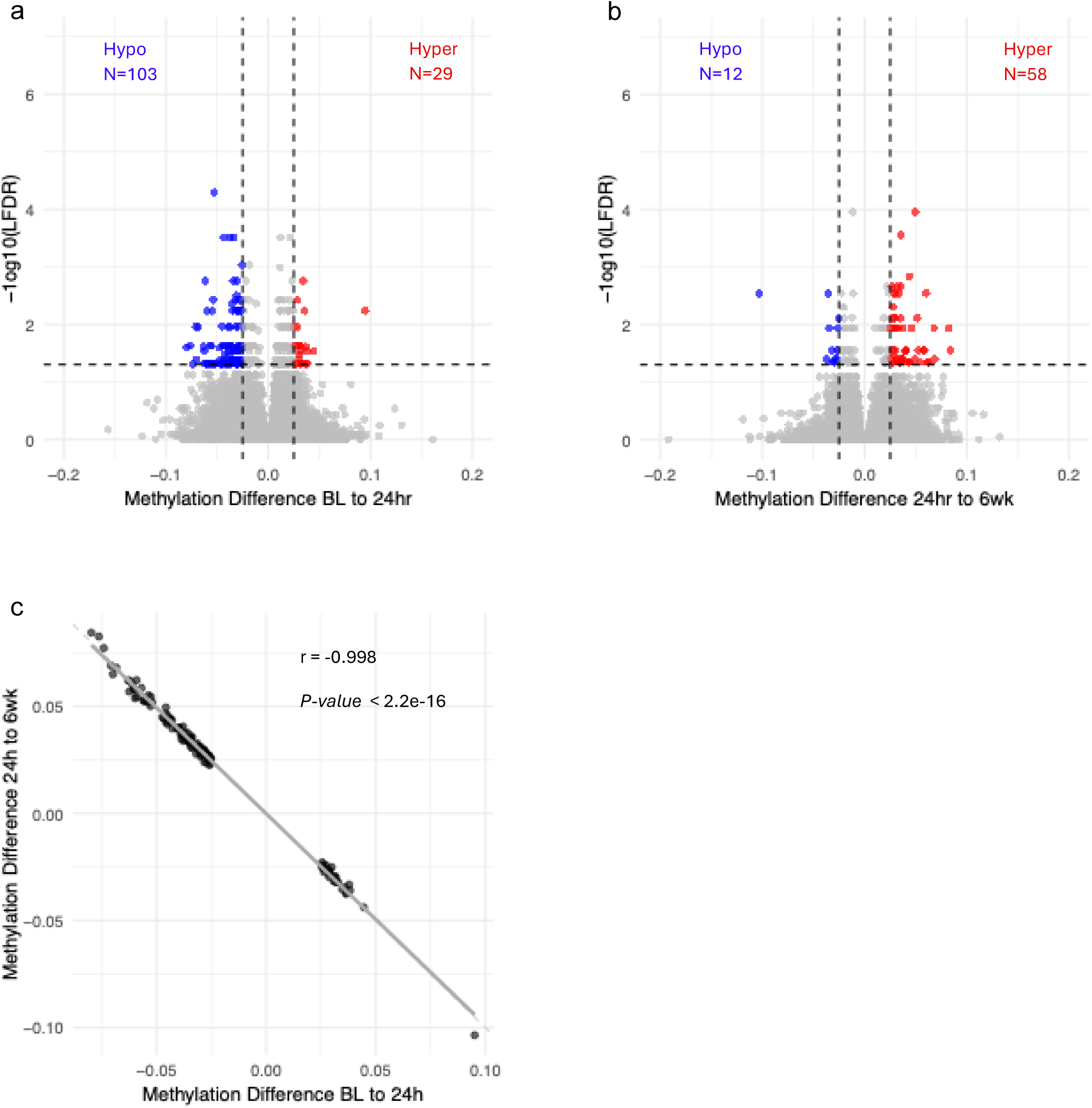
Differential DNA methylation levels between baseline, 24h and 6wk after anesthesia and surgery. Volcano plots show differentially methylated 5′-cytosine-phosphate-guanine-3′ (CpG) sites between (a) baseline (BL) and 24 hours after surgery (24h) comprising 103 hypomethylated and 29 hypermethylated differentially methylated positions (DMPs), and (b) 24h and 6 weeks after surgery (6wk) comprising 12 hypomethylated and 58 hypermethylated DMPs. Dashed vertical lines indicate a DNA methylation difference of 2.5% (x-axis). Dashed horizontal lines indicate a −log scale local false discovery rate (LFDR) of 0.05 (y-axis). Each point corresponds to a single CpG locus that is hypomethylated (blue), hypermethylated (red) or not differentially methylated (grey). (c) Scatter plot comparing the difference in methylation levels between BL and 24h (x-axis) *vs*. the difference in methylation levels between 24h and 6wk (y-axis). The line of best fit is depicted as a grey line. The regression of y = −1x is depicted as an overlapped grey dashed line visible at the top left and bottom right indicating that BL to 24h differentially methylated positions (DMPs) revert to BL in the 24h to 6wk comparison. Each point corresponds to one shared DMP in comparisons of BL and 24h and 24h and 6wk (N=135). The Pearson correlation coefficient is indicated by r.

### DNA methylation and postoperative delirium

Eight DMPs in 12 DMGs including *MBTD1*, *BID*, *PPAN, ANGPTL6*, PHF21B and *RBM5* were identified at baseline before surgery and anesthesia in participants who developed POD. (N=13) compared to participants without POD (N=81; Fig. 2a, Supplemental Table 3). Eighty-seven DMPs annotated to 91 DMGs were identified between participants with and without POD including *CLEC19A, FILIP1, ERICH1*, *PSENEN*, *SLC6A3*, and *TMEM196* in blood samples collected at 24h. (Fig. 2b, Supplemental Table 4). One DMP between participants with and without POD in samples collected at 6wk was identified in 2 genes, *FILIP1* and *LOC124901509* (Fig. 2c, Supplemental Table 5).

**Fig. 2.**
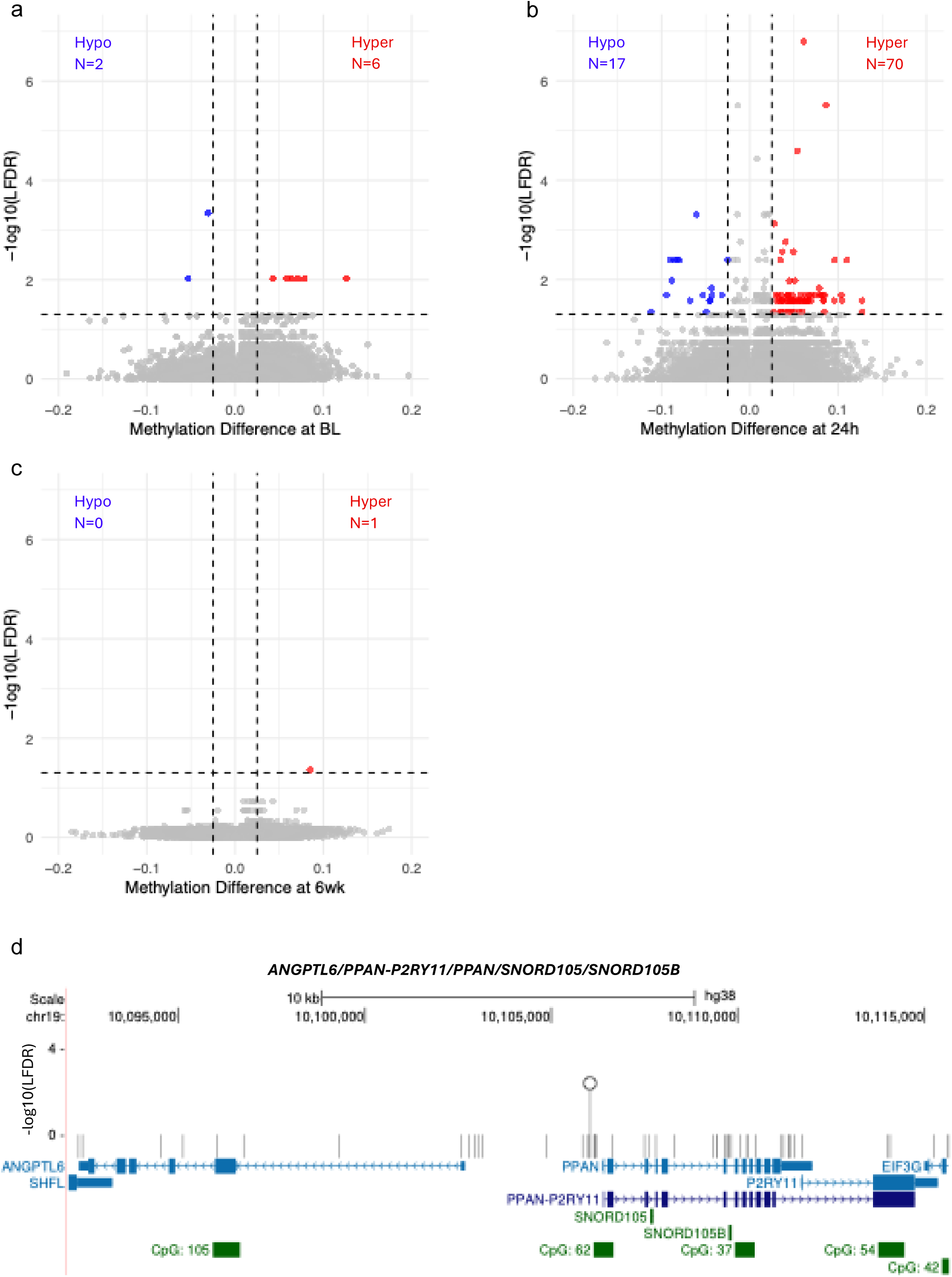
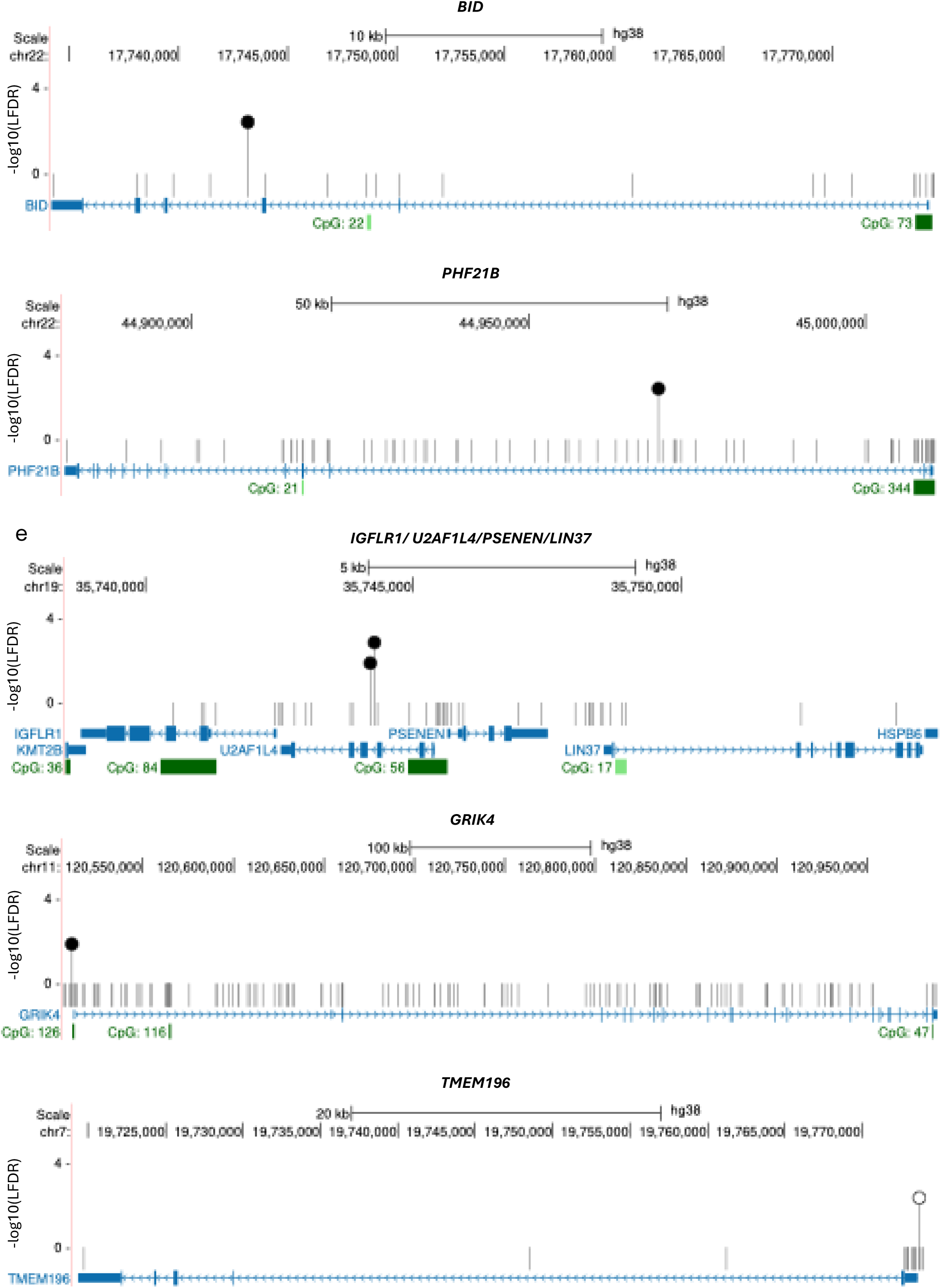
Differential methylation levels with and without postoperative delirium (POD) at 24h: Volcano plots show differentially methylated 5′-cytosine-phosphate-guanine-3′ (CpG) sites between (a) participants with and without postoperative delirium (POD) from samples acquired at baseline comprising 2 hypomethylated and 6 hypermethylated differentially methylated positions (DMPs), (b) participants with and without POD from samples acquired 24 hours after surgery (24h) comprising 17 hypomethylated and 70 hypermethylated DMPs, and (c) participants who developed POD and those who did not in samples acquired at 6 weeks (6wk) after surgery comprising 1 hypermethylated DMP. Dashed vertical lines indicate a DNA methylation difference of 2.5% (x-axis). Dashed horizontal lines indicate a −log scale local false discovery rate (LFDR) of 0.05 (y-axis). Each point corresponds to a single CpG locus that is hypomethylated (blue), hypermethylated (red) or not differentially methylated (grey). (d) University of California – Santa Cruz (UCSC) schematic diagrams of the methylome landscapes of genes with DMPs between participants with and without POD in samples acquired at baseline. Sense (>) and anti-sense (<) strands are depicted with gene symbols above each panel. Alignments to the human reference genome hg38 coordinates are displayed in base pairs (bp) at the top of each panel with the chromosome number (x-axis). Introns are depicted by thin blue lines and exons by thick blue boxes. CpG islands are indicated by green rectangles. The location of CpG dinucleotides in each gene is shown as a vertical line with a circle on top in white if hypomethylated or shaded in black if hypermethylated with a −log scale LFDR on the y-axis. One in 25 non-significant CpGs in each gene are shown by grey vertical tick marks.

### DNA methylation and postoperative neurocognitive disorder

No DMPs were identified between participants with and without PNCD at 6wk in samples acquired at baseline, 24h or 6wk.

## Discussion

Changes in DNA methylation levels after anesthesia and surgery and their roles in POD and PNCD are poorly characterized. In a cohort of 94 older participants who underwent general anesthesia and elective non-cardiac and non-neurologic surgeries, we identified 132 DMPs annotated to 198 DMGs between blood samples acquired at baseline before surgery and 24h afterwards (Supplemental Table 1). Genes comprising DMPs play critical roles in tissue regeneration, intracellular and immune signaling, RNA and DNA replication and stress responses, including the muscle-type acetylcholine receptor subunit *CHRNB1* (Gene ID: 1140, National Center for Biotechnology Information [NCBI], https://www.ncbi.nlm.nih.gov/) that encodes a subunit of the muscle acetylcholine receptor target of many anesthetics. The *LGALS1* gene (Gene ID: 3956) encodes a protein galectin-1 with a modulatory role in cell proliferation, differentiation, apoptosis and wound healing. The protein encoded by *SMAD4* (Gene ID: 4089) is activated by transforming growth factor (TGF)-beta signaling and is involved in signal cascade events in tissue regeneration. The SMAD4 protein participates in chondrogenesis, skeletal development and immune function. The gene encoding ryanodine receptor 2, *RYR2* (Gene ID: 6262) is expressed in cardiac muscle and is responsible for release of calcium ions from the sarcoplasmic reticulum. *CHST11* (Gene ID: 50515) encodes a protein that catalyzes the transfer of sulfate to chondroitin, a component of cartilage. *CDC25B* (Gene ID: 994) encodes a phosphatase that regulates entry into mitosis. *OBSCN* (Gene ID: 84033) belongs to a family of sarcomeric signaling proteins and is associated with cardiomyopathy and muscle organization. *ABHD16A* (Gene ID: 7920) encodes an enzyme that regulates lipid metabolism and intracellular signaling. As the primary brain phosphatidylserine hydrolase in the brain, ABHD16A is a critical regulator of immune signaling and lipid metabolism in neurons. DMPs observed at 24h reverted to baseline at 6wk in tandem with clinical recovery from anesthesia and surgery.

DMPs between samples collected before surgery and 24 hours after surgery were 78.03% hypomethylated most often associated with increased gene expression, and 21.97% hypermethylated most often associated with repressed gene expression. Caputi *et al*. used an antibody-based method to report global and gene-targeted DNA hypomethylation in samples collected during surgery from 11 patients undergoing general anesthesia and breast surgery. (Caputi, 2021) Global hypomethylation using an antibody-based method was confirmed by Li *et al*. in 24 patients sampled before and 7 days after surgery. (Li, 2020) Using an earlier Illumina microarray to interrogate 450,000 CpGs, Sadahiro *et al*. identified changes in DNA methylation associated with immune pathways in 55 participants on postoperative days 1 and 7. (Sadahiro, 2020) In samples collected on the third postoperative day, Bain *et al*. used the Illumina Infinium MethylationEPIC Beadchip to report hypermethylation on ribosomal RNA and protein metabolism pathways in 71 patients with extreme postoperative systemic inflammatory dysregulation. (Bain, 2022) The present data are the first to support longitudinal comparisons of DNA methylation levels in samples acquired at baseline before surgery and at 24h and 6wk thereafter, and the first to show that changes observed earlier in the perioperative interval return to BL at 6wk.

Eight DMPs in 12 DMGs distinguish participants at baseline before surgery who developed POD after surgery from those who did not. Representative baseline POD DMGs include *MBTD1*, a protein-coding gene (Gene ID: 54799) that serves as an epigenetic reader by recognizing methylated histone H4 to regulate chromatin structure and organization, gene transcription and DNA damage repair. The *BID* gene (Gene ID: 637) expresses a protein that acts both as a death agonist mediator of programmed cell death (apoptosis) and DNA damage repair. *PPAN* (Gene ID: 56342) encodes a nucleolar protein that is key for ribosomal biogenesis and mitochondrial homeostasis in support of cell proliferation, growth, and survival. The *ANGPTL6* protein (Gene ID: 83854) coordinates energy metabolism, promotes angiogenesis and blood flow in ischemic tissues, and supports tissue proliferation and regeneration. The protein encoded by *PHF21B* (Gene ID: 112885) also serves as an epigenetic reader in the brain that influences synaptic plasticity, regulates neurotransmission-related genes in the hippocampus, and directs cell fate toward neuronal differentiation during cortical development. *RBM5* (Gene ID: 10181) expresses a pre-mRNA splicing regulator that induces cell cycle arrest and apoptosis with activity at multiple target genes. Accordingly, DMPs in these DMGs compel replication and validation as biomarkers of POD susceptibility before surgery.

Eighty-seven DMPs in 91 DMGs distinguish participants with and without POD. Genes comprising DMPs with the most significant LFDRs play pivotal roles in intracellular signaling, neuronal migration, development and synaptic regulation, amyloid precursor protein processing and dopamine reuptake. For example, the *CLEC19A* protein (Gene ID: 100909498) is C-type lectin that binds carbohydrates involved in cell-to-cell communication and immune recognition. The *FILIP1* gene (Gene ID: 27145) encodes a protein that links the actin cytoskeleton and intracellular protein degradation and directs migration of neuronal cells from the ventricular zone during cortical development. *ERICH1* is a protein-coding gene (Gene ID: 157697) expressed in the cerebral cortex, amygdala, and cerebellum that participates in neuronal development, intracellular signaling and synaptic regulation. *PSENEN* (Gene ID: 55851) expresses a component of the γ-secretase complex that processes amyloid precursor protein implicated in Alzheimer’s disease pathogenesis. *SLC6A3* (Gene ID: 6531) encodes the dopamine transporter (DAT) protein that removes dopamine from the synapse for transport back into presynaptic neurons during neurotransmission. DAT is essential for termination of neural signals, regulation of dopamine levels in the brain, and supporting motivation, cognition, and movement. The cytoplasmic *TMEM196* protein (Gene ID: 256130) suppresses cellular proliferation and promotes apoptosis for removal of aging and damaged cells without harm to surrounding tissue. In 6wk samples from participants with and without POD, 1 DMP annotated to 2 genes, *FILIP1* (see above) and *LOC124901509*. *LOC124901509* (Gene ID: 124901507) encodes a small nucleolar U3 non-coding RNA that guides site-specific cleavage of ribosomal RNA (rRNA) during pre-rRNA processing. DMPs in these and multiple other DMGs that we report compel replication and validation as biomarkers of POD prevention, onset, progression, and intervention.

Earlier publications report an association between DNA methylation levels and POD using the Illumina Infinium MethylationEPIC Beadchip. Three studies examined patients undergoing neurosurgery without samples collected at the time of POD assessment. (Yamanashi, 2021, Wahba, 2022, Yamanashi, 2023). Two of 7 patients with POD in another study had dementia before surgery. (Nishizawa, 2024) Small sample sizes in the absence of prospective POD CAM testing by research personnel after neurosurgical procedures at the time of sample collection constrain comparisons of these reports with present data.

No DMPs were identified between participants with and without PNCD at 6wk in samples acquired at baseline, 24h and 6wk. It’s possible that PNCD is too subtle a central nervous system phenotype for detection with a molecular DNA methylation biomarker in circulating white blood cells. However, multiple investigations report a striking concordance between DNA methylation patterns in peripheral blood and brain tissue in participants with dementia and other central nervous system conditions. (Alberca, 2023, Zhang, 2024, Iturria-Medina, 2022, Silva, 2022, Wei, 2020, Karlsson, 2023, Stevenson, 2022, Mendonca, 2024, Horvath, 2012) PNCD was first recognized as a distinct research construct 150 years after the introduction of general anesthesia, and PNCD case reports outside the research setting have yet to appear. The PNCD phenotype may be minimally disruptive to the human methylome compared to changes seen with more pronounced neurodegenerative conditions. In turn, the Illumina Infinium MethylationEPIC Beadchip interrogates < 4% of all CpG loci in the human genome that have been selected by the manufacturer to reside in CpG islands and promoters as preferred commercial targets for cancer and related phenotypes, rather than comprehensive 100% coverage of > 25,000,000 CpGs identifiable by whole genome methylation sequencing (WGMS) as we’ve recently reported. (Madrid, 2025)

Our study has multiple strengths. Criteria for testing positive with POD and PNCD are formally validated, consistent with contemporary research analytic standards and conducted by trained staff with linkage between the time of cognitive testing and sample acquisition. An effect size threshold of at least 2.5% meets and exceeds multiple reports of differential DNA methylation levels measured across diverse human phenotypes. (Breton, 2017, Perzel Mandell, 2021, Campagna, 2021, Min, 2021, Christensen, 2024, Abrishamcar, 2024) Prior studies using the Illumina Infinium MethylationEPIC Beadchip report significant findings in similarly sized and smaller data sets. (Ham, 2019, Mordaunt, 2019, Rizardi, 2019, Grimm, 2019, Papale, 2019) Corrections for multiple comparisons and repeated measure were conservative and uniformly applied.

Our study must also be interpreted in keeping with several limitations. The MADCO-PC and INTUIT studies used different instruments for detecting delirium (*i.e*., CAM *vs*. 3D-CAM) that may have increased variance in the relationship strength seen in differential DNA methylation levels between the two cohorts. Of note, both instruments provide high sensitivity and specificity for detecting delirium, and both methods were supplemented by delirium chart reviews to minimize missed cases. Though moderate the delirium rate (12.6%) was comparable to that reported in other studies of similarly aged non-cardiac surgical participants. (Schenning, 2015) In addition to the absence of a no-surgery cohort, participants with pertinent demographic characteristics and predisposing factors were not included or balanced including, for example, participants with divergent POD and PNCD severities, intercurrent medications, and types of surgery and types of anesthesia. (Dilmen, 2023)

On the strength of our findings using the EPIC array, future investigations comprising whole genome methylation sequencing data are clearly warranted for detection of new biomarkers of neurocognitive changes after anesthesia and surgery together with individualized whole genome sequencing for identification methylation quantitative trait loci (meQTLs) (Smith, 2014), transcriptomics, and plasma and CSF proteomics. Enrollment of a greater number of participants with POD is a high priority enabled by multicenter integration of ongoing POD protocols, and inclusion of participants who differ by ancestry, age, co-existing disorders, and pre-existing signs of cognitive decline arising from Alzheimer’s disease and related disorders.. Larger cohorts will support more precise effect estimates for the strength of association between differential DNA methylation, POD and PNCD, will test potential interaction effects between additional baseline factors and perioperative risks, and comprise analyses with continuous as well as dichotomous variables as potential indicators of severity.

The present data support the design of future studies to test the validity of a panel of DMPs and DMGs as biomarkers of POD risk in minimally invasive blood samples configured for rapid translation to clinical and research applications. In addition to epigenetically primed responses that arise after surgery and anesthesia, differential DNA methylation is sensitive to environmental exposures and conditions that increase POD risk including mild cognitive impairment and Alzheimer’s disease. Accordingly, DNA methylation biomarkers offer a promising new tool for understanding POD pathogenesis and for delirium risk stratification.

## Data Availability

All data produced in the present study are available upon reasonable request to the authors.

## Details of authors’ contributions

Study concept and design: KH, RA, MB

Statistical analysis planning: RA, AM, SK

Acquisition and analysis of data: KH, MB, SK, AM, BH, MW, MD, MS, RA

Data pre-processing: RA, AM

Principle investigator for INTUIT: MB

Principle investigator for MADCO-PC: MB

Manuscript writing: KH, RA and MB with assistance from MD, AM, SK and BH

Subject matter expertise in project planning and manuscript refinement: KH, MD, RA, MB

All authors provided critical feedback on the paper

## Acknowledgements

The authors thank the participants and study personnel who made this work possible.

## Appendix A. Supplementary data

Supplementary data to this article may be found online at:

## Data archived at

GEO accession number:

---

**Supplementary Fig. 1.**
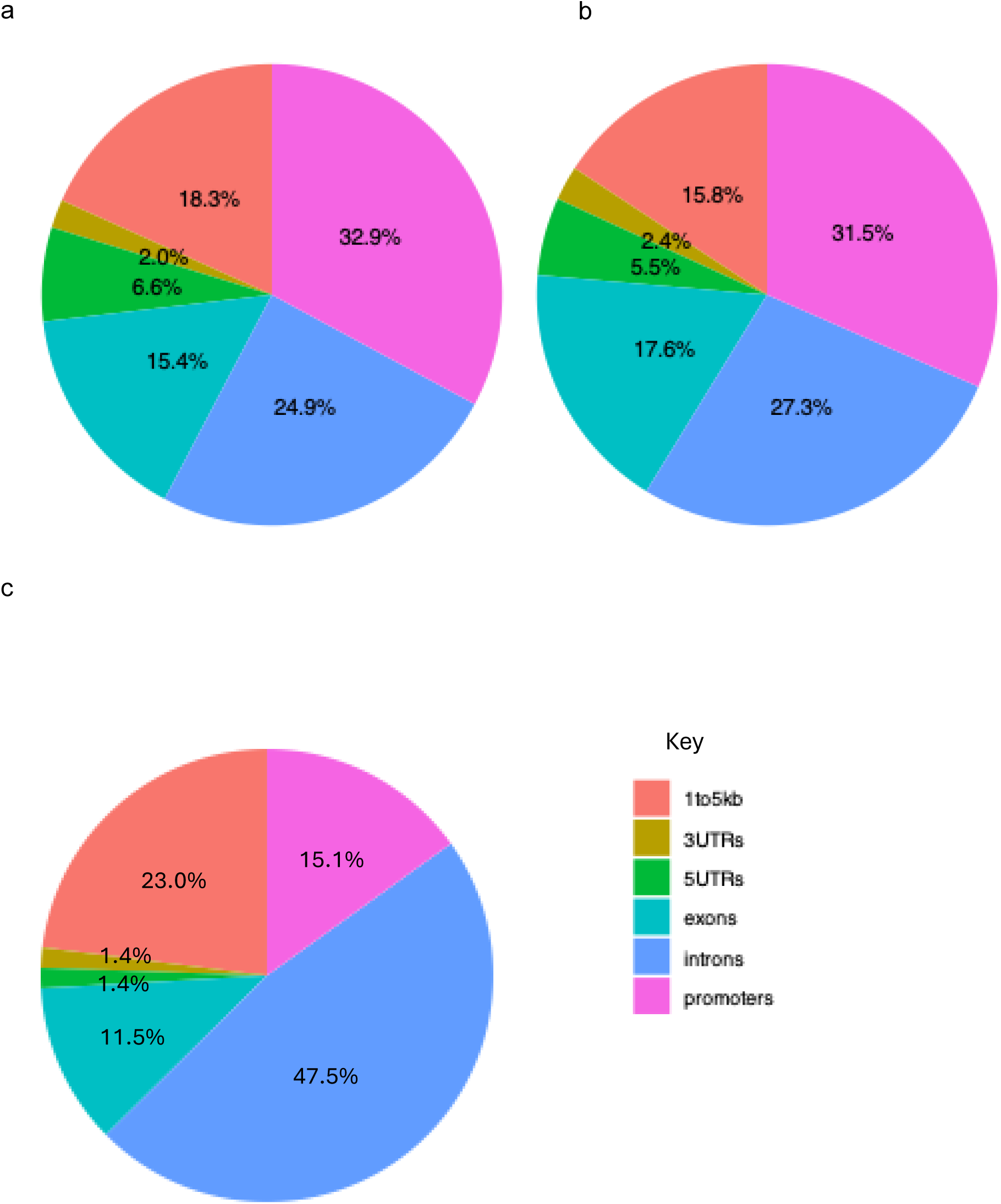
Differentially methylated positions (DMPs) annotated to gene structures including introns, exons, 5’ untranslated region (5UTR), 3’ untranslated region (3UTR), 1 to 5 kilobases upstream of the transcription start site (TSS) (1to5kb), and 1kb upstream of the TSS (promoter) between (a) samples acquired at baseline to samples acquired 24h after surgery (N=132 DMPs), (b) samples acquired at 24h after surgery to 6wk after surgery (N=70 DMPs), and (c) samples acquired at 24h from participants with and without POD (N=87 DMPs).

